# Long COVID-19 in Hospitalized and Ambulatory Children in Jamaica

**DOI:** 10.1101/2025.09.29.25335807

**Authors:** Samantha Ann-Patrice Mosha Miller, Mark Daniel Hicar, Crista-Lee Shahine Berry, Kathryn B. Anderson, John F. Lindo, Gene D. Morse, Celia Dana Claire Christie

## Abstract

A retrospective observational cohort study evaluated acute illness predictors of pediatric Long COVID in Jamaica. Poisson regression modeling associated acute univariate symptoms of vomiting, abdominal pain, ageusia, anosmia, fatigue, confusion, brain fog, and multisystem inflammatory syndrome in children (MIS-C); multivariable composite predictors of MIS-C, vomiting, and ageusia. A high incidence in our cohort supports needs for improvements applicable in resource-constrained settings.

## INTRODUCTION

The COVID-19 pandemic effect on Jamaican children was similar to other centers.^**1**^ The initial striking post-COVID-19 condition included those who developed “Multi-system Inflammatory Syndrome in Children” (MIS-C), with hyperinflammation involving ≥2 organ systems occurring 2-6 weeks after acute SARS-CoV-2 infection.^**2**^ Child and adolescent acute COVID-19 case rates ranged from 0.60% to 16.9%, in 15 Caribbean countries/territories compared with a global prevalence of 20.2% in 2021.^**3**^ While globally, persistence of symptoms, termed “Long COVID-19”, was observed in as high as 61.9% of children and adolescents.^**4**^

Despite these early studies, COVID-19 in Caribbean countries is understudied; in particular, later findings of Long COVID. We evaluated the risk factors and long-term impact of COVID-19 in children and adolescents aged <16 years with acute COVID-19 who were hospitalized or received ambulatory care from January 2021 through December 2023 at the University Hospital of the West Indies (UHWI), in the middle-developing, Afro-Caribbean, island-nation of Jamaica. We defined the circulation of the Delta SARS-CoV-2 “Variant of Concern” as those identified during 2021 and Omicron in those occurring from 2022 through 2023, referencing Jamaica’s national surveillance criteria.

## METHODS

This retrospective observational cohort study used the UHWI’s laboratory data management system to locate all positive SARS-CoV-2 PCR’s, rapid antigen, and serological tests for patients aged <16 years. Positive laboratory listings were cross-referenced with UHWI’s emergency room and hospital inpatient registers, and pediatric infectious disease consultations. A modified WHO COVID-19 case report form was used to extract data from inpatient and outpatient hospital charts. We then conducted oral telephone interviews using a modified International Severe Acute Respiratory and Emerging Infection Consortium (ISARIC) COVID-19 long-term follow-up survey. These data were evaluated to determine those who met the WHO 2023 definition for Long COVID-19 in children and adolescents and to assess their outcome.^**5**^ Ethical approval for the study was obtained from the Mona Campus Research Ethics Committee, of the University of the West Indies.

All main variables had less than 5% missingness, which is considered negligible by statistical guidelines. A robust Poisson regression model was used with the Huber/White/sandwich estimator to account for variable follow-up times (median 24 months with interquartile range, IQR: 18-36), and robust standard errors corrected for over-dispersion.^**6**^ The models produced incidence rate ratios based on clinical and demographic factors. Both univariate and multivariable models were generated, with the latter assessed using Pseudo R^2^, Akaike Information Criterion (AIC), and Bayesian Information Criterion (BIC). Pearson’s chi-squared, Wilcoxon rank-sum, and Kruskal-Wallis tests evaluated differences in the distribution of clinical and demographic factors by hospitalization status and MIS-C diagnosis. Analyses of functional impairment and quality-of-life were conducted using ordinal logistic regression, with these measures as the outcomes. All regression models included follow-up time as a covariate. Statistical significance was set at p<0.05.

## RESULTS

During the 3 years, the UHWI laboratory system reported 704 positive SARS-CoV-2 records for patients aged <16 years. Seventy-four participants were interviewed from 177 who had medical records available and verified contact information (**Supplemental Figure 1**). All were Afro-Caribbean, with 54.1% (n=40) males and 45.9% (n=34) females with a mean age of 9.1 ± 5.6 years.

Among acute COVID-19 infections, the Delta SARS-CoV-2 variant accounted for 40.5% (n=30) and Omicron for 59.5% (44) (**Supplemental Table 1**). Laboratory-confirmed SARS-CoV-2 infection was present in 94.5% (n=69); 63.8% (n=44) had positive PCR results, 29.0% (n=20) had positive antigen test results, and 30.4% (n=21) were diagnosed serologically, with some overlap among those groups. In Jamaica, COVID-19 vaccines were only offered to children aged ≥12 years. Six children (8.1%) were vaccinated. Of the 27 patients ≥12 years, 22% received a vaccine.

Our main outcomes were Long COVID-19, MIS-C, and hospitalization (**Supplemental Table 1**). Almost half of our subjects (n=35;47.3%) met WHO criteria for Long COVID-19. Twenty-four (32.4%) developed MIS-C. Another 32.4% (n=24) were hospitalized with acute COVID-19 and 20% developed Long COVID-19. Chronic illnesses included sickle cell disease, asthma, obesity, neurological disorders, and food allergies (**Supplemental Table 1**). The most frequently reported acute COVID-19 symptoms were fever (55.4%), cough (43.2%), shortness of breath (21.6%), fatigue (17.6%), rhinorrhea (16.2%), vomiting (14.9%), headache (10.8%), and ageusia (9.5%). Several symptoms demonstrated statistically significant associations with Long COVID-19 based on the univariate robust Poisson regression models (**Figure**). Regarding fatigue, 25.7% of those who went on to develop Long COVID-19 experienced this symptom, compared to 10.3% who did not develop Long COVID-19 (IRR 1.84; 95% CI 1.26-2.68; p=0.002). Significant differences were also noted with vomiting (25.7% vs 5.1%; IRR 2.18; 95% CI 1.39-3.41; p=0.001), diarrhea (14.3% vs 2.6%; IRR 2.03; 95% CI 1.12-3.69; p=0.020), anosmia (11.4% vs 0.0%; IRR 1.65; 95% CI 1.24-2.18; p = 0.001), abdominal pain (5.7% vs 0.0%; IRR 1.78; 95% CI 1.21-2.59; p=0.003), confusion (5.7% vs 0.0%; IRR 2.56; 95% CI 1.34-4.90; p=0.004), brain fog (5.7% vs 0.0%; IRR 1.78; 95% CI 1.38-2.29; p=0.000) (**Figure**).

**Figure.**
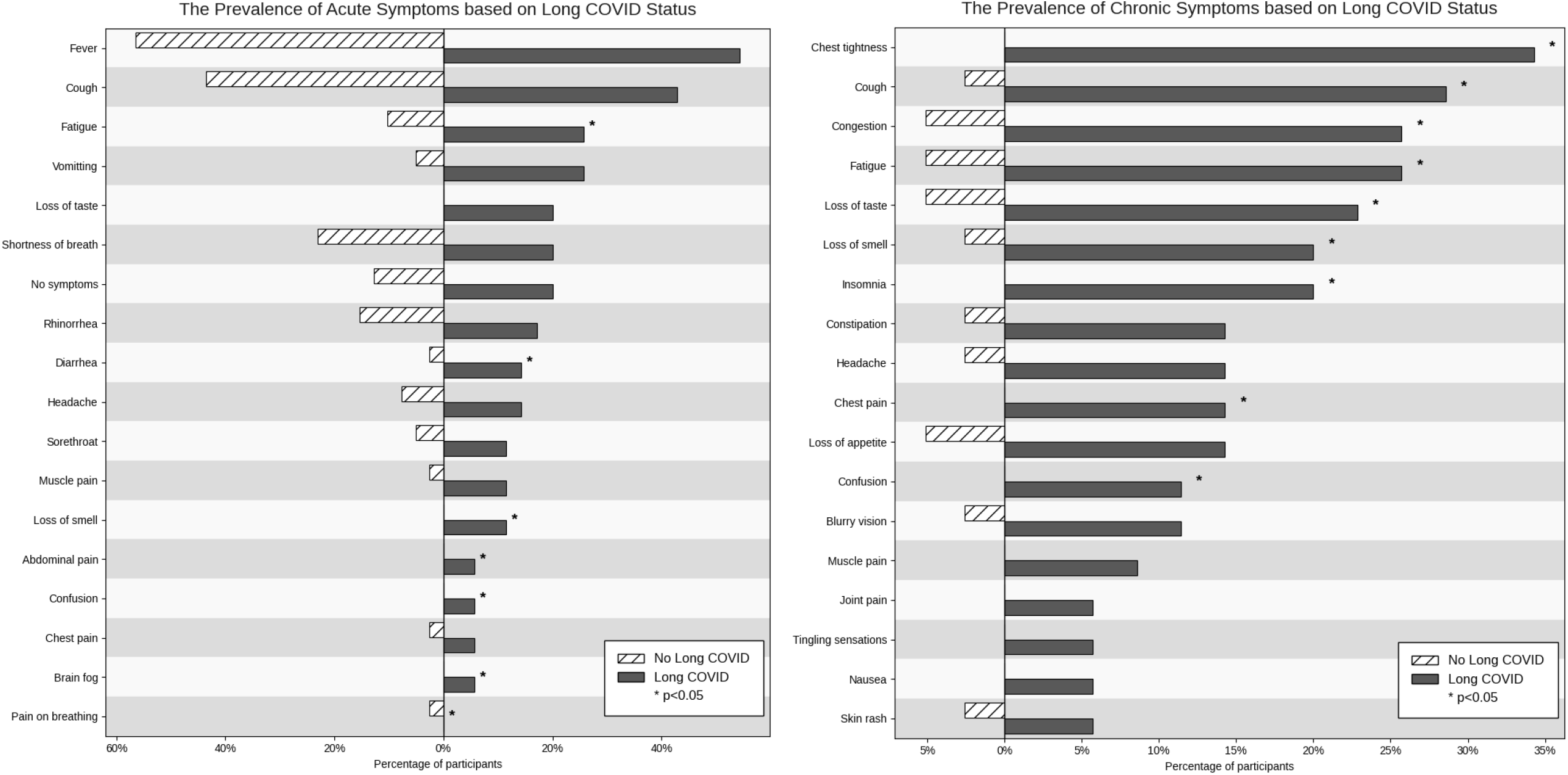
Prevalence of Acute and Chronic Symptoms based on Long-COVID Status.

The most common persistent symptoms were cough (85.1%), chest tightness (16.2%), fatigue (14.9%), congestion (14.9%), ageusia (13.5%), anosmia (10.8%), anorexia (9.5%), and insomnia (9.5%). All these symptoms and chest pain (6.8%) showed a statistically significant association with the development of Long COVID-19. Chest tightness was experienced by 34.3% of those who met the criteria for Long COVID-19 vs. 0.0% of those who did not (IRR 2.81; 95% CI 1.96-4.02; p=0.000). Significant results for the remaining symptoms were: cough 28.6% vs 2.6% (IRR 0.37; 95% CI 0.25-0.54; p=0.000); nasal congestion 25.7% vs 5.1% (IRR 2.34; 95% CI 1.60-3.42; p=0.000), fatigue 25.7% vs 5.1% (IRR 1.77; 95% CI 1.13-2.79; p=0.013); ageusia 22.9% vs 5.1% (IRR 1.61; 95% CI 1.04-2.51; p=0.034); insomnia 20.0% vs 0.0% (IRR 2.15; 95% CI 1.53-3.04; p=0.000); anosmia 87.5% vs 12.5% (IRR 1.85; 95% CI 1.27-2.71; p=0.001); and chest pain 14.3% vs 0.0% (IRR 2.56; 95% CI 1.89-3.47; p=0.000) (**Figure**).

Functional impairment was assessed based on mobility, self-care, school attendance, pain or discomfort, and negative emotions before and after the COVID-19 infection. Most participants reported having no problems in any domain, regardless of whether they developed Long COVID-19 (**Supplemental Table 2**). Quality of life was also assessed through changes in activities, such as eating, sleeping, neuropsychiatric symptoms, social relationships, and emotional well-being, with no significant associations observed.

We investigated the relationship between elevated inflammatory markers on presentation to the hospital and the development of Long COVID-19 (**Supplemental Table 3**). Although C-reactive protein, erythrocyte sedimentation rate, D-dimer, lactate dehydrogenase, and absolute neutrophil counts were all higher in the Long COVID-19 group, these differences were not statistically significant. Surprisingly, ferritin levels were significantly elevated in the non-Long COVID-19 group. Our hospitalized MIS-C cohort received several interventions, including anticoagulants, anti-inflammatory drugs, intravenous immunoglobulin (IVIG), aspirin, antivirals, antibiotics, and corticosteroids, without significant association between individual interventions and the development of Long COVID-19 (**Supplemental Table 3**).

Univariate Poisson regression models revealed single predictors of the development of Long COVID-19 (**Supplemental Table 4**). Significant associations were found in patients with MIS-C as well as several acute COVID-19 symptoms, including confusion, fatigue, vomiting, abdominal pain, anosmia, ageusia, and brain fog (p<0.05; IRR>1.0). Multivariate analysis demonstrated that MIS-C, vomiting, and ageusia together formed a composite predictive model for Long COVID-19 (Pseudo R^2^:11.9%; likelihood ratio test: p<0.001) (**Supplemental Table 4**).

## DISCUSSION

The long-term impact of COVID-19 was assessed using the WHO 2023 Pediatric Long COVID-19 criteria. Our 47.3% prevalence of pediatric Long COVID-19 synchronizes with the 20.0%-61.9% in the global literature.^**4**,**8**^ but our study design would add significant recall and selection bias among those without symptoms. The most common acute COVID-19 symptoms were fever, cough, shortness of breath, fatigue, and rhinorrhea, with fatigue and vomiting being significant risk factors for Long COVID-19.

Our study mirrored persistent Long COVID-19 symptoms of cough, chest tightness, fatigue, congestion, ageusia, anosmia, anorexia, and insomnia.^**4**,**8-10**^ The RECOVER study also cited gastrointestinal, neuropsychiatric, and pain-related complaints as predictors in school-aged children, while adolescents experiencing ageusia, anosmia, fatigue, and pain were more likely to develop Long COVID-19.^**10**^

There were no significant associations between Long COVID-19 and demographics, hospitalization status, or circulating SARS-CoV-2 variants, although our study lacked direct sequencing, so strains were estimated based on community circulation. Surprisingly, acute ferritin levels were significantly higher in those who did not develop Long COVID-19.

There were no statistically significant impairments of function or quality of life in children with Long COVID-19. Individual predictors of Long COVID-19 were acute COVID-19 symptoms, including vomiting, abdominal pain, ageusia, anosmia, fatigue, confusion, brain fog, and MIS-C. Composite predictors were MIS-C, vomiting, and ageusia. The RECOVER study similarly noted the predictive value of neuropsychiatric symptoms.^**10**^ Long COVID-19 may therefore increase the mental health burden on children, adolescents, and the healthcare and educational system if it is not addressed.

## CONCLUSION

Half of our identified cohort met WHO 2023 criteria for Pediatric Long COVID-19.^**5**^ This would require increased resources for robust SARS-CoV-2 testing and pharmacotherapy, which may be inaccessible in resource-constrained settings.

## Data Availability

All available data are posted in the manuscript.

