## Supplemental tables and figure for "Long COVID-19 in Hospitalized and Ambulatory Children in Jamaica"

**Supplemental Table 1: Distribution of baseline clinical and demographic characteristics among Jamaican children being evaluated for Long COVID**

| Characteristics | Total Sample | Hospitalized (n=24, 32.4%) | Not hospitalized (n=50, 67.6%) | MIS-C present (n = 20, 27.0%) | MIS-C absent (n=54, 73.0%) | Long COVID present (n=35, 47.3%) | Long COVID absent (n=39, 52.7%) |
|---|---|---|---|---|---|---|---|
| **Gender** | | | | | | | |
| Male | 40, 54.1% | 13 (54.2%) | 27 (54.0%) | 11 (55.0%) | 29 (53.7%) | 16 (45.7%) | 24 (61.5%) |
| Female | 34, 45.9% | 11 (45.8%) | 23 (46.0%) | 9 (45.0%) | 25 (46.3%) | 19 (54.3%) | 15 (38.5%) |
| **Variant** | | | | | | | |
| Delta | 30, 40.5% | 8 (33.3%) | 22 (44.0%) | 10 (50.0%) | 20 (37.0%) | 17 (48.6%) | 13 (33.3%) |
| Omicron | 44, 59.5% | 16 (66.7%) | 28 (56.0%) | 10 (50.0%) | 34 (63.0%) | 18 (51.4%) | 26 (66.7%) |
| **Age groups** | | | | | | | |
| < 6 yrs | 31, 41.9% | 8 (33.3%) | 23 (46.0%) | 7 (35.0%) | 24 (44.4%) | 11 (31.4%) | 20 (51.3%) |
| 6-12 yrs | 16, 21.6% | 6 (25.0%) | 10 (20.0%) | 8 (40.0%) | 8 (14.8%) | 9 (25.7%) | 7 (18.0%) |
| > 12 yrs | 27, 36.5% | 10 (41.7%) | 17 (34.0%) | 5 (25.0%) | 22 (40.7%) | 15 (42.9%) | 12 (30.8%) |
| **Diagnosis** | | | | | | | |
| Lab confirmed | 69, 94.5% | 21 (91.3%) | 48 (96.0%) | 16 (84.2%) | 53 (98.2%) | 33 (97.5%) | 36 (94.7%) |
| Physician confirmed | 3, 4.1% | 1 (4.4%) | 2 (4.0%) | 2 (10.5%) | 1 (1.9%) | 1 (2.6%) | 1 (2.6%) |
| Self-diagnosed | 1, 1.4% | 1 (4.4%) | 0 (0.0%) | 1 (5.3%) | 0 (0%) | 1 (2.6%) | 1 (2.6%) |
| **Antigen positive** | 20, 34.5% | 6 (28.6%) | 14 (37.8%) | 2 (13.3%) | 18 (41.8%) | 8 (28.6%) | 12 (40.0%) |
| **Antigen negative** | 38, 65.5% | 15 (71.4%) | 23 (62.2%) | 13 (86.7%) | 25 (58.1%) | 20 (71.4%) | 18 (60.0%) |
| **PCR positive** | 44, 59.5% | 13 (54.2%) | 31 (62.0%) | 0 (0.0%) | 44 (81.5%) | 19 (54.3%) | 25 (64.1%) |
| **PCR negative** | 30, 40.5% | 11 (45.8%) | 19 (38.0%) | 20 (100.0%) | 10 (18.5%) | 16 (45.7%) | 14 (35.9%) |
| **Serology positive** | 21, 48.8% | 6 (54.6%) | 15 (46.9%) | 16 (88.9%) | 5 (20.0%) | 12 (54.6%) | 9 (42.9%) |
| **Serology negative** | 22, 51.2% | 5 (45.5%) | 17 (53.1%) | 2 (11.1%) | 20 (80.0%) | 10 (45.5%) | 12 (57.1%) |
| **Asthma** | | | | | | | |
| Present | 17 (23.0%) | 4 (16.7%) | 13 (26.0%) | 5 (25.0%) | 12 (22.2%) | 9 (25.7%) | 8 (20.5%) |
| Absent | 57 (77.0%) | 20 (83.3%) | 37 (74.0%) | 15 (75.0%) | 42 (77.8%) | 26 (74.3%) | 31 (79.5%) |
| **Obesity** | | | | | | | |
| Present | 9 (12.2%) | 2 (8.3%) | 7 (14.05) | 3 (15.0%) | 6 (11.1%) | 4 (11.4%) | 5 (12.8%) |
| Absent | 65 (87.8%) | 22 (91.7%) | 43 (86.0%) | 17 (85.0%) | 48 (88.9%) | 31 (88.6%) | 34 (87.2%) |
| **Neurological disorders** | | | | | | | |
| Present | 7 (9.5%) | 3 (12.5%) | 4 (8.05) | 20 (100.0%) | 7 (13.0%) | 3 (8.6%) | 4 (10.3%) |
| Absent | 67 (90.5%) | 21 (87.5%) | 46 (92.0%) | 0 (0.0%) | 47 (87.0%) | 32 (91.4%) | 35 (89.7%) |
| **History of Food Allergies** | | | | | | | |
| Present | 7 (9.5%) | 2 (8.3%) | 5 (10.0%) | 2 (10.0%) | 49 (90.7%) | 3 (8.6%) | 4 (10.3%) |
| Absent | 67 (90.5%) | 22 (91.7%) | 45 (90.0%) | 18 (90.0%) | 5 (9.3%) | 32 (91.4%) | 35 (89.7%) |
| **Sickle cell disease** | | | | | | | |
| Present | 17 (23.0%) | 2 (8.3%) | 5 (10.0%) | ***5 (25.0%)** | ***2 (3.7%)** | 3 (8.6%) | 4 (10.3%) |
| Absent | 57 (77.0%) | 22 (91.7%) | 45 (90.0%) | ***15 (75.0%)** | ***52 (96.3%)** | 32 (91.4%) | 35 (89.7%) |

† $p<0.05$: * and bold;

**Supplemental Table 2. Quality of life and functional impairment among Jamaican children being evaluated for Long COVID**

| Health parameter | Total (n=74) | | Long COVID Present (n=35, 47.3%) | | Long COVID Absent (n=39, 52.7%) | | P-value |
|---|---|---|---|---|---|---|---|
| | Initial | Follow-up | Initial | Follow-up | Initial | Follow-up | |
| **Mobility** | | | | | | | 0.000 |
| No problems | 39, 97.5% | 37, 92.5% | 21, 95.5% | 20, 90.9% | 18, 100.0% | 17, 94.4% | |
| Some problems | 0, 0.0% | 1, 2.5% | 0, 0.0% | 1, 4.6% | 0, 0.0% | 0, 0.0% | |
| Many problems | 1, 2.5% | 2, 5.0% | 1, 2.6% | 1, 4.6% | 0, 0.0% | 1, 5.6% | |
| **Self-care** | | | | | | | 0.237 |
| No problems | 36, 90.0% | 36, 90.0% | 21, 95.5% | 21, 95.5% | 15, 83.3% | 15, 83.3% | |
| Some problems | 1, 2.5% | 2, 5.0% | 0, 0.0% | 0, 0.0% | 1, 5.6% | 2, 11.1% | |
| Many problems | 3, 7.5% | 2, 5.0% | 1, 4.6% | 1, 4.6% | 2, 11.1% | 1, 5.6% | |
| **Usual activities** | | | | | | | 0.155 |
| No problems | 37, 92.5% | 35, 87.5% | 20, 90.9% | 19, 86.3% | 17, 94.4% | 16, 88.9% | |
| Some problems | 2, 5.0% | 3, 7.5% | 1, 4.6% | 2, 9.1% | 1, 5.6% | 1, 5.6% | |
| Many problems | 1, 2.5% | 2, 5.0% | 1, 4.6% | 1, 4.6% | 0, 0.0% | 1, 5.6% | |
| **Pain or discomfort** | | | | | | | 0.997 |
| No problems | 28, 70.0% | 24, 60.0% | 14, 63.6% | 12, 54.6% | 14, 77.8% | 12, 66.7% | |
| Some problems | 8, 20.0% | 15, 37.5% | 6, 27.3% | 9, 40.9% | 2, 11.1% | 6, 33.3% | |
| Many problems | 4, 10.0% | 1, 2.5% | 2, 9.1% | 1, 4.6% | 2, 11.1% | 0, 0.0% | |
| **Negative emotions** | | | | | | | 0.631 |
| No problems | 25, 62.5% | 20, 50.0% | 15, 68.2% | 11, 50.0% | 10, 55.6% | 9, 50.0% | |
| Some problems | 13, 32.5% | 18, 45.0% | 6, 27.3% | 10, 45.5% | 7, 38.9% | 8, 44.4% | |
| Many problems | 2, 5.0% | 2, 5.0% | 1, 4.6% | 1, 4.6% | 1, 5.6% | 1, 5.6% | |

**Supplemental Table 3. Inflammatory markers and Therapeutic Interventions in Jamaican children being evaluated for Long COVID**

| | **Long COVID-19 Present (n=35, 47.3%)** | **Long COVID-19 Absent (n=39, 52.7%)** |
|---|---|---|
| **Inflammatory Markers** | **Median (interquartile range)** | **Median (interquartile range)** |
| C-Reactive Protein | 4.4 (0.6-5.5) | 3.9 (0.5-4.4) |
| Erythrocyte Sedimentation Rate | 98.5 (21-130) | 29.5 (11-65) |
| D-dimer | 2245 (497-4936) | 1407 (247-2154) |
| **Ferritin** | ***265.5 (143-1520)** | ***337.5 (155-464)** |
| Lactate Dehydrogenase | 485 (354-682) | 440 (326-715) |
| Absolute Neutrophil Count | 10.7 (3.3-65) | 9.2 (2.8-23.9) |
| **Interventions** | **Frequency (%)** | **Frequency (%)** |
| Anti-inflammatory drugs<br>Present<br>Absent | 1, 2.9%<br>34, 97.1% | 1, 2.6%<br>38, 97.4% |
| Immunoglobulin<br>Present<br>Absent | 6, 17.1%<br>29, 82.9% | 4, 10.3%<br>35, 89.7% |
| Aspirin<br>Present<br>Absent | 5, 14.3%<br>30, 85.7% | 3, 7.7%<br>36, 92.3% |
| Antivirals<br>Present<br>Absent | 1, 2.9%<br>34, 97.1% | 2, 5.1%<br>37, 94.9% |
| Antibiotics<br>Present<br>Absent | 13, 37.1%<br>22, 62.9% | 18, 46.2%<br>21, 53.9% |
| Corticosteroids<br>Present<br>Absent | 9, 25.7%<br>26, 74.3% | 12, 30.8%<br>27, 69.2% |

† $p<0.05$: * and bold; † Reference ranges: C-Reactive Protein: < 1.0 mg/dL; Erythrocyte Sedimentation Rate < 20 mm/hr; D-dimer < 500 ng/mL; Ferritin: 10-330 ng/mL; Lactate Dehydrogenase:105-200 U/L; Absolute Neutrophil Count:1.63-6.96

**Table 4. Univariate and Multivariate Poisson Regression assessing incidence of Long COVID among Jamaican children**

| | Uni-variables | | | | Multi-variables | | | |
|---|---|---|---|---|---|---|---|---|
| **Predictors** | **Incidence Rate Ratio (IRR)** | **95% Confidence Interval** | **Robust Standard Error** | **P-value** | **Incidence Rate Ratio (IRR)** | **95% Confidence Interval** | **Robust Standard Error** | **P-value** |
| **Age groups** (ref. less than 6 years) | | | | | | | | |
| **6-12 years** | 1.6 | 0.8, 3.0 | 0.5 | 0.16 | 1.5 | 0.8, 2.8 | 0.5 | 0.24 |
| **12 years and older** | 1.6 | 0.9, 2.8 | 0.5 | 0.13 | 1.4 | 0.7, 2.6 | 0.4 | 0.35 |
| **Female** (ref: Male) | 1.4 | 0.9, 2.3 | 0.4 | 0.18 | 1.5 | 0.9, 2.6 | 0.4 | 0.13 |
| **Hospitalized** (ref: not hospitalized) | 0.5 | 0.3, 1.0 | 0.2 | 0.06 | 0.6 | 0.3, 1.1 | 0.2 | 0.08 |
| **Omicron** (ref: Delta) | 0.7 | 0.5, 1.2 | 0.2 | 0.18 | 0.8 | 0.5, 1.3 | 0.2 | 0.32 |
| **MIS-C present** (ref: absent) | 1.6 | 1.0, 2.5 | 0.4 | 0.046 | 1.8 | 1.0, 3.0 | 0.5 | 0.03 |
| **Confusion** (ref: absent) | 2.2 | 1.7, 2.8 | 0.3 | <0.001 | | | | |
| **Fatigue** (ref: absent) | 1.6 | 1.0, 2.6 | 0.4 | 0.042 | 1.2 | 0.7, 2.2 | 0.4 | 0.45 |
| **Vomiting** (ref: absent) | 2.0 | 1.3, 3.0 | 0.4 | 0.001 | 2.7 | 1.6, 4.4 | 0.7 | <0.001 |
| **Abdominal pain** (ref: absent) | 2.2 | 1.7, 2.8 | 0.3 | <0.001 | | | | |
| **Loss of smell** (ref: absent) | 2.3 | 1.7, 2.9 | 0.3 | <0.001 | 0.5 | 0.2, 1.2 | 0.2 | 0.14 |
| **Loss of taste** (ref: absent) | 2.3 | 1.8, 3.2 | 0.4 | <0.001 | 3.2 | 1.7, 5.9 | 1.0 | <0.001 |
| **Brain fog** (ref: absent) | 2.2 | 1.7, 2.8 | 0.3 | <0.001 | | | | |

† 'ref:' is an abbreviation for 'reference:'

† Empty cells in the multivariate table represent independent variables that did not fit into the best fitting multivariate model

### Supplemental Figure: Flow chart of Participants

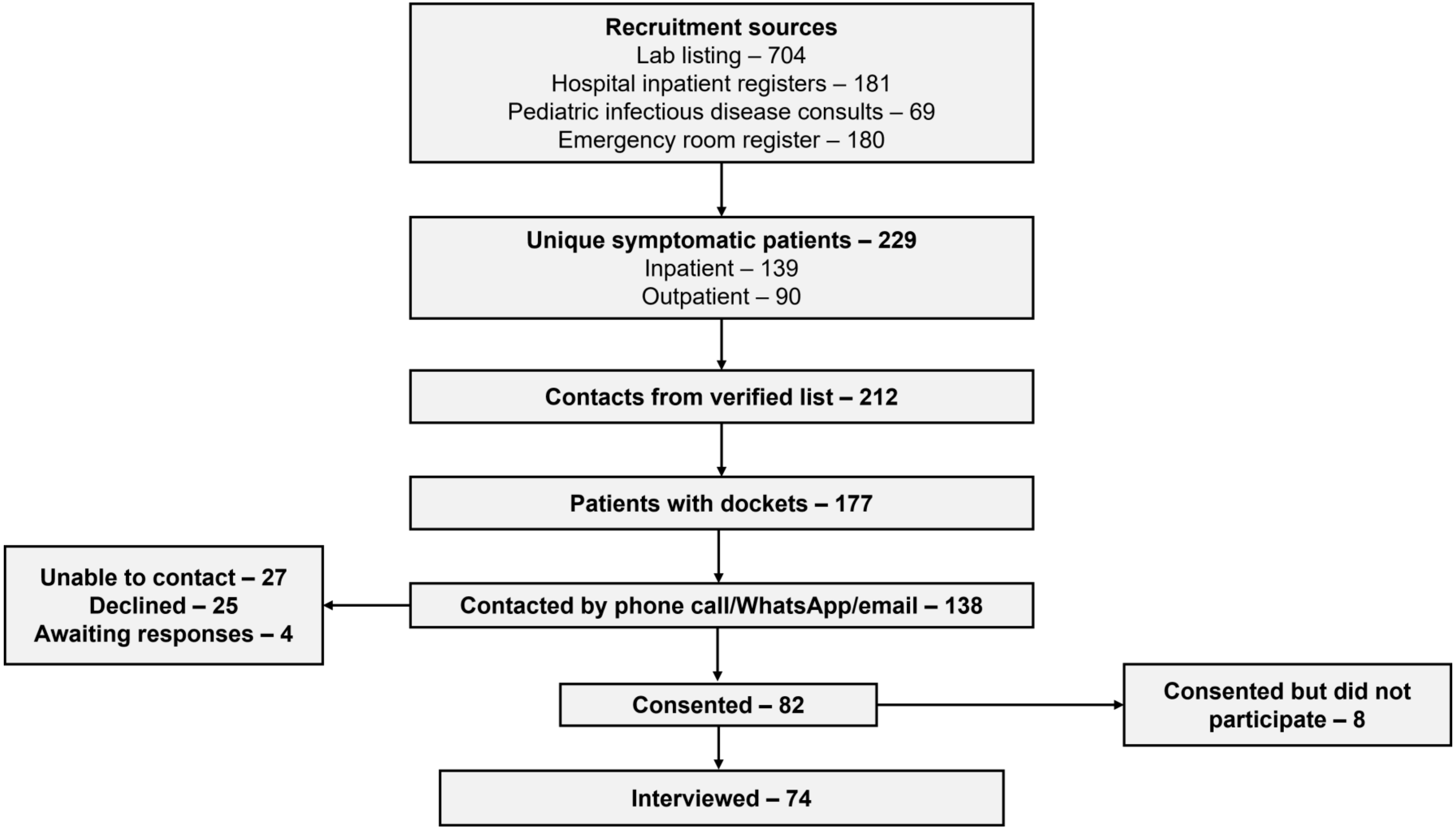
